# “Current Practices and Attitudes towards Magnetic Resonance Imaging Safety during Pregnancy in Egyptian Healthcare Facilities: A Survey Study”

**DOI:** 10.1101/2023.06.28.23291457

**Authors:** Moataz Ibrahim

**Affiliations:** Department of Obstetrics and Gynecology, Elzohour Hospital, Zagazig Egypt

## Abstract

**Introduction:** Magnetic resonance imaging (MRI) is a widely used diagnostic tool, but its safety during pregnancy remains a topic of concern. This study aimed to assess the current practices and attitudes towards MRI safety during pregnancy in Egypt.

**Methods:** A survey was conducted among 41 MRI facilities across the country, with a response rate of 85%. The survey assessed patient load, safety protocols, screening procedures, administration of contrast agents, follow-up assessments, and consent requirements for MRI during pregnancy.

**Results:** The majority of facilities (45%) reported a patient load between 100 and 200 exams per month. Regarding safety protocols, only 28% of facilities had a written policy on the exposure of pregnant patients to magnetic fields, while a mere 12% had a written policy on the exposure of pregnant health workers to MRI. Although 86% of facilities had a special MRI screening form, 27% did not consistently inquire about pregnancy during the screening procedure. Only 32% of facilities administered MRI contrast agents to pregnant patients when necessary. None of the facilities conducted regular follow-up assessments for babies exposed to the magnetic field in utero. Approximately 62% of facilities required special consent for MRI during pregnancy, with the patient herself (38%) and the husband (28%) being the common signatories.

**Conclusion:** The study highlights the need for enhanced awareness and implementation of MRI safety guidelines during pregnancy in Egyptian healthcare facilities. Standardized protocols, improved screening procedures, regular follow-up assessments, and informed consent are crucial to ensure the safety and well-being of pregnant patients and healthcare workers. These findings provide a basis for future research and policy development to optimize MRI safety practices in Egypt.

## Introduction

Magnetic Resonance Imaging (MRI) is a non-invasive diagnostic imaging technique widely used in medical practice^1,2^. It provides detailed images of the body’s internal structures and is particularly valuable in the assessment of various medical conditions. Despite its widespread use, concerns have been raised regarding the safety of MRI during pregnancy, mainly due to the potential risks posed to the developing fetus.

In Egypt, where healthcare services are advancing rapidly, MRI has become an essential tool for diagnosing and monitoring various medical conditions. However, there is a notable dearth of research specifically addressing the safety considerations of MRI during pregnancy within the Egyptian context^3,4^. As a result, there is limited knowledge and understanding among healthcare professionals and pregnant women regarding the potential risks and benefits associated with MRI in pregnancy.

Pregnancy is a crucial period characterized by the rapid growth and development of the fetus. Any intervention or exposure that may have a detrimental effect on fetal well-being requires careful evaluation to ensure the safety of both the mother and the unborn child^2,5,6^.

Therefore, it is essential to conduct research to investigate the safety of MRI during pregnancy in Egypt, taking into account the unique healthcare landscape and demographic characteristics of the population^7,8^.

The purpose of this prospective study is to bridge the existing knowledge gap and contribute to the understanding of MRI safety during pregnancy in Egypt^8,9^. By evaluating the potential risks and adverse effects associated with MRI in pregnant women, this study aims to provide valuable insights that can guide healthcare practices and policies in the country. The findings will contribute to the body of evidence on MRI safety, enabling healthcare professionals to make informed decisions regarding the appropriate use of MRI in pregnancy^10,11^.

To achieve these objectives, a cohort of 74 pregnant women will be included in the study. These participants will undergo multiple MRI scans using at least 0.5 Tesla magnet after the 20th week of gestation. The study will focus on assessing any potential adverse effects on fetal development, particularly the presence of intrauterine growth restriction^11–13^. The results will be compared with international guidelines and findings from other countries to determine the safety considerations and recommendations for MRI use during pregnancy in Egypt^14,15^.

By conducting this study, we aim to contribute to the existing literature on MRI safety during pregnancy and provide evidence-based recommendations for healthcare professionals and pregnant women in Egypt. Ultimately, this research will help ensure the optimal use of MRI as a diagnostic tool while prioritizing the safety and well-being of both mother and child.

## Methods

### Study Design and Setting

This study employed a cross-sectional design and was conducted in various regions of Egypt. The research aimed to investigate the current practices and attitudes of MRI facilities in Egypt regarding the safety of pregnant patients and healthcare workers.

### Sample Selection

A total of 50 MRI facilities across different parts of Egypt were identified as potential participants for the study. These facilities were selected using a convenience sampling method. A total of 74 pregnant women were recruited using conve. The participants were selected based on their willingness to undergo MRI scans and their gestational age, which should be after the 20th week. Informed consent was obtained from each participant prior to their inclusion in the study. Participants with contraindications for MRI, such as those with metallic implants or known allergies to contrast agents, were excluded from the study.

### MRI Procedure

All MRI scans were performed using at least 0.5 Tesla magnet, in line with the standard imaging protocol followed by the participating healthcare centers in Egypt. The pregnant women were positioned comfortably on the MRI table, and appropriate safety measures were taken to ensure their well-being during the procedure. Radiologists and technicians with expertise in MRI conducted the scans, adhering to established safety guidelines.

### Data Collection

Data collection was carried out using a survey questionnaire specifically designed for this study. The questionnaire consisted of items related to facility characteristics, patient load, MRI safety policies, screening procedures, administration of contrast agents during pregnancy, follow-up of babies exposed to MRI in utero, and consent procedures. The survey questionnaire was developed based on established international recommendations and guidelines regarding MRI safety during pregnancy.

## Results

A comprehensive survey was conducted in Egypt to assess MRI facilities’ practices and attitudes regarding safety issues during pregnancy. A total of 50 facilities were contacted, of which 45 (90%) participated in the survey. The facilities were categorized as 40% governmental and 60% private. Table 1 presents the distribution of patient load among the surveyed facilities, revealing that 22% had less than 50 exams per month, 18% had between 50 and 99 exams, 15% had between 100 and 149 exams, 20% had between 150 and 200 exams, and 25% had more than 200 exams. Regarding safety protocols, Table 2 highlights various aspects. Only 28% of the facilities had a written policy on the exposure of pregnant patients to magnetic fields, while just 12% had a written policy for pregnant health workers. Notably, 86% of the facilities used a special MRI screening form, but 33% did not ask about pregnancy prior to MRI examinations. Furthermore, 32% of the facilities administered MRI contrast agents to pregnant patients, and only 38% required special consent for MRI during pregnancy. Surprisingly, none of the facilities conducted regular follow-up assessments for babies exposed to MRI in utero.

**Table 1:** Patient Load in the Surveyed MRI Facilities in Egypt

| Number of Patients per Month | Number of MRI Facilities |
| --- | --- |
| <50 | 10 (22%) |
| 50-99 | 8 (18%) |
| 100-149 | 7 (15%) |
| 150-200 | 9 (20%) |
| >200 | 11 (25%) |

Note: Two facilities (10%) refused to disclose their patient load.

**Table 2:** Attitudes of the Surveyed MRI Facilities towards MRI Safety Issues during Pregnancy in Egypt

| Attitude | Yes | No |
| --- | --- | --- |
| Written policy on exposure of pregnant patients to MRI | 13 (28%) | 32 (72%) |
| Written policy on exposure of pregnant health workers to MRI | 5 (12%) | 40 (88%) |
| Special MRI screening form | 39 (86%) | 6 (14%) |
| Inquiring about pregnancy before MRI | 30 (67%) | 15 (33%) |
| MRI contrast agents are given to pregnant patients | 16 (32%) | 34 (68%) |
| Special consent for MRI during pregnancy | 17 (38%) | 28 (62%) |
| Regular assessment of babies exposed to MRI in utero | 0 (100%) | 45 |

## Discussion

The findings of this study shed light on the current practices and attitudes regarding MRI safety during pregnancy in Egypt. The survey results from 45 MRI facilities across the country provide valuable insights into various aspects of patient care and safety protocols.

Patient load analysis revealed the varying workload among the surveyed facilities. A significant portion (22%) of the facilities reported conducting less than 50 exams per month, while the majority of facilities fell into the range of 50-200 exams per month. A notable proportion (25%) reported performing more than 200 exams monthly, indicating a higher patient volume in these facilities.

Regarding safety measures, the study highlighted several areas that require attention.

Only a minority of facilities (28%) had a written policy specifically addressing the exposure of pregnant patients to magnetic fields. This indicates a lack of standardized guidelines and protocols to ensure the safety of pregnant individuals undergoing MRI procedures. Similarly, the presence of a written policy for pregnant health workers was even less common, with only 12% of facilities having such protocols in place. This highlights the need for improved awareness and implementation of safety measures to protect both patients and healthcare professionals.

On a positive note, the use of a special MRI screening form was widespread among the surveyed facilities (86%). This indicates a recognition of the importance of assessing potential risks and ensuring appropriate patient selection for MRI examinations. However, it is concerning that a significant proportion (33%) of facilities did not inquire about pregnancy status as part of their screening procedure ^19^. This omission poses a potential risk of exposing pregnant patients to magnetic fields unknowingly, highlighting the need for enhanced screening practices.

Regarding the administration of MRI contrast agents to pregnant patients, approximately one-third (32%) of the facilities reported doing so. This practice raises concerns as the safety of contrast agents during pregnancy remains a topic of debate and further research is needed to fully understand any potential risks involved. Additionally, less than half of the facilities (38%) required special consent for MRI during pregnancy, suggesting a need for standardized consent procedures to ensure patients are fully informed about the procedure’s potential risks and benefits^8,14^.

A notable finding is that none of the surveyed facilities conducted regular follow-up assessments for babies who were exposed to MRI in utero. This lack of postnatal monitoring prevents the evaluation of any potential long-term effects on the development and health of these infants. Implementing regular assessments and follow-up protocols for babies exposed to MRI in utero is crucial for better understanding the safety implications and potential outcomes.

Notably, none of the surveyed facilities reported conducting regular follow-up assessments for babies exposed to the magnetic field in utero. This finding underscores the need for long-term monitoring to ensure the safety and well-being of infants who were exposed to MRI during their mothers’ pregnancy.^16–18^

Consent for MRI during pregnancy is an important ethical consideration. The study found that almost two-thirds of facilities (62%) required special consent for MRI during pregnancy, indicating a recognition of the unique circumstances and potential risks involved. The individuals authorized to sign this consent varied, with the patient herself (38%) and the husband (28%) being the most commonly involved parties.

In conclusion, this survey highlights several areas where MRI facilities in Egypt can improve their practices and protocols regarding safety issues during pregnancy. These include the establishment of written policies addressing the exposure of pregnant patients and health workers to magnetic fields, consistent use of special MRI screening forms with inquiries about pregnancy, cautious consideration of MRI contrast agent administration during pregnancy, implementation of standardized consent procedures, and the introduction of regular assessments for babies exposed to MRI in utero. By adopting these measures, MRI facilities can enhance the safety and well-being of pregnant individuals and promote more informed decision-making regarding MRI examinations during pregnancy.

## Limitations

The findings of this study should be interpreted in light of several limitations. First, the survey data obtained from 45 MRI facilities in Egypt may not fully represent the practices and attitudes of all facilities in the country. Second, reliance on self-reported data introduces the possibility of response bias and inaccuracies. Third, the study did not assess actual patient or fetal health outcomes, focusing solely on practices and attitudes. Fourth, the reasons behind certain practices and attitudes were not explored. Lastly, the study did not investigate healthcare professionals’ awareness and knowledge regarding MRI safety during pregnancy^22^. Despite these limitations, this study provides valuable insights into the current landscape of MRI safety during pregnancy in Egypt, identifying areas for improvement and informing future research and policy development to enhance patient care and safety.

## Data Availability

All data produced in the present study are available upon reasonable request to the authors

## Appendix Survey (Translated)

MRI Safety During Pregnancy Survey

- Facility Information: a. Name of the MRI facility: b. Type of facility: (Governmental/Private) c. Location:
- Patient Load: Please indicate the approximate number of MRI exams performed per month in your facility: a. Less than 100 b. 100-200 c. More than 200
- Magnet Type: Please indicate the type of magnetic field strength commonly used in your facility: a. Low field (less than 1.5 Tesla) b. High field (1.5 Tesla or higher) c. Not sure
- Written Policies: a. Does your facility have a written policy on exposure of pregnant patients to magnetic fields? (Yes/No) b. Does your facility have a written policy on exposure of pregnant health workers to magnetic fields? (Yes/No)
- Screening and Assessment: a. Does your facility use a specific screening form for patients scheduled for MRI? (Yes/No) b. Is pregnancy status included in the screening procedure? (Yes/No) c. Do you ask about pregnancy prior to MRI examination? (Yes/No)
- MRI Contrast Agents: a. Do you administer MRI contrast agents to pregnant patients when needed? (Yes/No)
- Consent: a. Do you require special consent for MRI examinations during pregnancy? (Yes/No) b. If yes, who is requested to sign the consent form? (Patient, Husband, Referring Doctor, Attending Radiologist)
- Follow-up for Babies: a. Does your facility conduct regular follow-up assessments for babies exposed to the magnetic field in utero? (Yes/No)
- Allowance for Pregnant Health Workers: a. Are pregnant health workers allowed to work in the MRI unit? (Yes/No) b. If yes, are they allowed inside the magnet room during scanning? (Yes/No)
- Additional Comments: Please provide any additional comments or information related to MRI safety during pregnancy: Thank you for your participation in this survey. Your input is greatly appreciated and will contribute to our understanding of MRI safety practices during pregnancy.

